# Cytosponge procedures produce fewer respiratory aerosols and droplets than oesophagogastro-duodenoscopies

**DOI:** 10.1101/2023.05.31.23290803

**Authors:** George S.D. Gordon, Samantha Warburton, Sian Parkes, Abigail Kerridge, Adolfo Parra-Blanco, Jacobo Ortiz-Fernandez-Sordo, Rebecca C. Fitzgerald

**Author notes:** **AUTHOR CONTRIBUTIONS** *GSDG*: Study design, statistical analysis of data, manuscript preparation. *SW, SP*: Data collection and curation, patient consent. *AK*: Study design, ethics. *APB*: Study concept and design. *JOFS*: Study concept and design, study supervision. *RFC*: Study concept and design, data analysis, manuscript preparation. **DATA AVAILABILITY STATEMENT** Data associated with this publication is available at http://dx.doi.org/10.17639/nott.XXX Code used for data analysis in this publication can be found at https://github.com/gsdgordon/aerosols. **FUNDING** The authors also thank Norgine Pharmaceuticals for sponsoring the purchase of a particle counter. GSDG acknowledges a UKRI Future Leaders Fellowship (MR/T041951/1).

## Abstract

**Background and Aims:** Oesophagogastro duodenoscopies (OGD) are aerosol generating procedures (AGP) that may spread respiratory pathogens. We aim to investigate production of airborne aerosols and droplets during Cytosponge procedures, which are being evaluated in large-scale research studies and NHS implementation pilots to reduce endoscopy backlogs.

**Design and Methods:** We measured 18 Cytosponge and 37 OGD procedures using a particle counter (diameters 0.3μm-25μm) taking measurements 10cm from the mouth. Two particle count analyses were performed: whole procedure and event-based.

**Results:** Direct comparison with duration-standardised OGD procedures shows Cytosponge procedures produce 2.16x reduction (*p*<0.001) for aerosols and no significant change for droplets (*p=*0.332). Event-based analysis shows particle production is driven by throat spray (aerosols:138.1x reference, droplets:16.2x), which is optional, and removal of Cytosponge (aerosols:14.6x, droplets:62.6x). Cytosponge coughing produces less aerosols than OGD (2.82x, *p*<0.05).

**Conclusions:** Cytosponge procedures produce significantly less aerosols and droplets than OGD procedures and thus reduce two potential transmission routes for respiratory viruses.

## Background

It is well-established that OGD procedures produce aerosols and droplets (defined as particles ≤5μm and > 5μm in diameter respectively) and are thus aerosol generating procedures (AGP) ^1–3^. Aerosols can remain airborne for many hours before depositing in the lower airways, whereas droplets land quickly and can contaminate surfaces: these two size ranges therefore represent two key routes of transmission for respiratory viruses such as a SARS-CoV-2. OGD procedures therefore present significant occupational risk to healthcare workers, necessitating the use of mitigation strategies including use of high-grade personal protective equipment ^4^, improved ventilation ^5,6^, increased fallow periods ^7^ and alternative procedures (e.g. transnasal endoscopy) ^1^. Whilst these are all effective to some degree, they have significant downsides including incurring significant cost and medical waste, and increased time per patient leading to backlogs.

A Cytosponge procedure involves the patient swallowing a capsule on a string, which dissolves in the stomach to release a sponge that collects cells from the oesophagus as it is pulled out. Cytosponge can replace some OGD procedures, are effective in detection and monitoring of Barrett’s oesophagus and also substantially cheaper than OGDs as they can be administered by a single nurse in an office setting^8,9^. During COVID-19, Cytosponge procedures have been implemented in pilots across NHS England and NHS Scotland for patients with reflux symptoms referred for a routine endoscopy, and for patients undergoing Barrett’s surveillance. However, while it has been assumed they are less aerosol generating than OGD due to the nature of the procedure, which does not require continual flushing and suction, their aerosol generating potential has never been measured. In this study we use a previously validated methodology for measuring aerosols and droplets in OGD procedures and apply this to Cytosponge procedures^1^.

## Methods

The methodology for this observational study is based on a previous ‘baseline’ study of aerosol generation in digestive endoscopy ^1^. The OGD arm (Wales Ethics Committee IRAS no. 285595) included patients undergoing routine upper GI endoscopy at Nottingham University Hospitals (NUH) NHS Trust between October 2020-March 2021. The Cytosponge arm (England REC IRAS no. 283505, amendment 3) included patients undergoing Cytosponge procedures at NUH NHS Trust between September 2022-February 2023. Informed consent was obtained for all participants.

Particle counts were measured and analysed using an AeroTrak particle counter (TSI, Shoreview MN, model 9500-01) with an isokinetic inlet head placed 10cm from the patient’s mouth via a 2m tube (manufacturer provided, length calibrated). The particle counter measures particle counts in six diameter ranges (0.5-0.7μm, 0.7-1.0μm, 1.0-3.0μm, 3.0-5.0μm, 5.0-10.0μm, 10.0-25μm) and has a flow rate of 100L/min, with readings averaged over 7s (the minimum permitted by the instrument). All staff in the room wore masks (surgical or FFP3) to minimise the contribution of additional human aerosol sources.

For whole procedure analysis, we first normalize particle counts for procedure length to create an effective count for a 20 minute procedure. We next identify a 5-minute reference window before the procedure starts to use for statistical comparison. To minimise impact of slowly-varying room particle background, we perform a second analysis in which a median filter is used to subtract this background leaving behind only sharp increases (‘spikes’) in particle counts. This neglects slow increases in the room background caused by continuous patient respiration and so is provided alongside a comparison of raw particle counts.

Aerosol-producing events are analysed using a background subtraction approach described in our previous methodology ^1^. Specifically, we consider the following individual aerosol generating events: insertion of Cytosponge, removal of Cytosponge, application of anaesthetic throat spray and coughing/gagging during procedure. The insertion and removal of Cytosponge are compared against intubation and extubation events for OGD procedures. Cytosponge procedure events are compared both against a ‘null reference’ event in which no activity occurs and against similar events in the OGD group.

All statistical analysis was performed using MATLAB software (The MathWorks Inc., Massachusetts). Building on existing models of respiratory aerosol production we model particle counts using a log-normal distribution and can therefore apply a *t-*test to logarithmically transformed data. For individual events the data distribution is modelled as the sum of a log-normal and normal distribution to account for negative particle counts arising from the subtraction step. A boot-strapping method provides numerical estimates of *p*-values between events.

## Results

The demographic data for the two groups of patients is given in Table 1. No variables are significantly different except for the use of anaesthetic Xylocaine throat spray: this was used for 100% of OGD patients, but only 22% of Cytosponge patients according to patient preference. Within the OGD group our previously published analysis found no significant effect of midazolam on aerosol or droplet production^1^.

**Table 1:** Patient demographics

| Variable | Study group | OGD | Cytosponge |
| --- | --- | --- | --- |
| **n** |  | 37 | 18 |
| **Age** |  | Range: 24-93<br>Median: 61 | Range: 40-78<br>Median: 67<br>(*p*=0.541) |
| **Sex** |  | Male: 23, Female: 14 | Male: 16, Female: 2<br>(*p*=0.142) |
| **BMI** |  | Range: 16.3-38.2<br>Median: 24.8 | Range: 22.7-34.3<br>Median: 27.0<br>(*p*=0.077) |
| **Smoking** |  | Smoker: 9<br>Non-smoker: 28 | Smoker: 2<br>Non-smoker: 16<br>(*p*=0.441) |
| **Sedation** |  | Midazolam + throat spray: 16<br>Throat spray only: 21 | No sedation: 14<br>Throat spray only: 4<br>(*p* < 0.001) |
| **No. coughing/gagging events per 20 mins** |  | Mean: 1.97 | Mean: 1.46<br>(*p*=0.379) |
| **Procedure duration (minutes)** |  | Mean: 7.2 | Mean: 8.1<br>(*p*=0.247) |

For the full Cytosponge procedure analysis, for particles in the aerosol size range we find there is no significant difference with the reference (i.e. no procedure) window (*p*=0.083) and similarly for particles in the droplet size range (*p=*0.940). However, using the background subtraction approach we find that there are 3.7x more particles produced in the aerosol size range (95% CI: 1.91x – 7.22x, *p<*0.001) and 2.2x more particles produced in the droplet size range (95% CI: 1.29x – 3.73x, *p<*0.01) compare to reference window.

Next, we directly compare particle production in Cytosponge procedures vs. standard OGD (Figure 1a). In the aerosol size range Cytosponge produces 2.16x fewer particles per unit time than OGD (95% CI: 1.48x – 3.13x, *p*<0.001) but for the droplet size range there is no significant difference (*p=*0.332). When comparing only against Cytosponge procedures where no anaesthetic throat spray is administered, we find in the aerosol size range Cytosponge procedures produces 2.08x fewer particle per unit time than OGD (95% CI: 1.38x – 3.13x, *p*<0.001) and in the droplet size range there is no significant different (*p=*0.693). Further, when applying the background subtraction we find that Cytosponge produces 4.39x fewer aerosols per unit time than OGD (95% CI 2.41x – 8.02x, *p*<0.001) and 2.23x fewer droplets (95% CI 1.34x – 3.71x, *p*<0.01).

**Figure 1:**
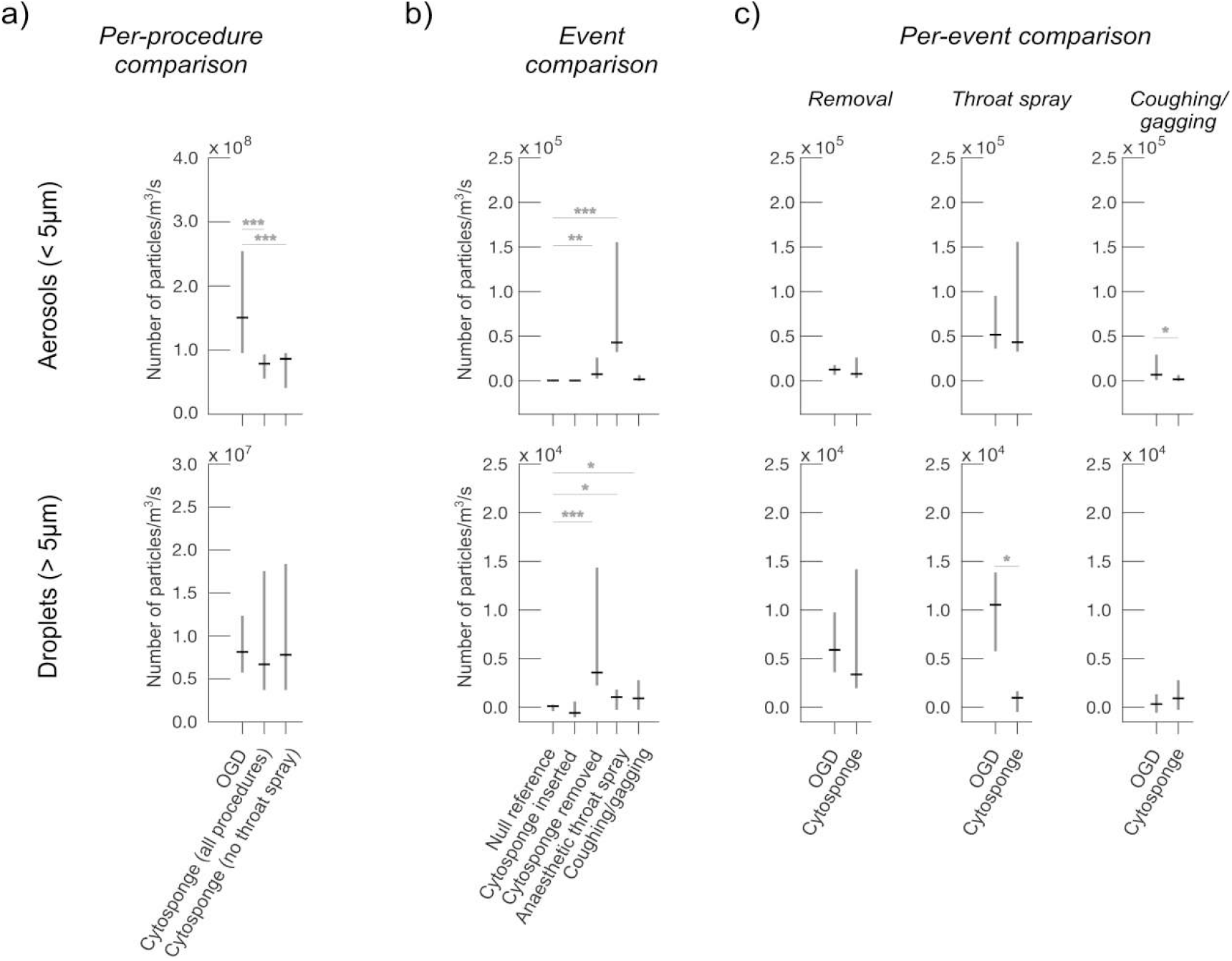
a) Comparison between OGD and Cytosponge (both including and excluding anaesthetic throat spray) whole-procedure particle counts. b) Comparison between events within Cytosponge procedures, indicating statistically significant production of particles. c) Comparison between similar events in OGD and Cytosponge procedures. Insertion is excluded because it is not significantly particle producing in either OGD or Cytosponge. *p<0.05, **p<0.01, ***p<0.001.

We next compare events during Cytosponge procedures (Figure 1b.). In the aerosol size range, the statistically significant events are Cytosponge removal (14.6x null reference, 95% CI: 1.80x – 242.3x, *p*<0.01, n=17) and application of throat spray (138.1x, 95% CI: 13.9x – 2713x, p < 0.001, n=4). Cytosponge insertion was not significant (*p*=0.420) nor was coughing/gagging (*p*=0.112). In the droplet size range, the statistically significant events are Cytosponge removal (62.6x null reference, 95% CI: 6.7x – 1476x, *p*<0.01, n=17), application of throat spray (16.2x, 95% CI: 0.58x - 442.5x, *p<*0.05, n=4) and coughing/gagging (14.6x null reference, 95%CI: 0.8x – 369.5x, *p<*0.05). Cytosponge insertion was not significant (*p=*0.434)

Finally, we compare statistically significant equivalent events from OGD and Cytosponge procedures. In the aerosol size range we find coughing/gagging produces 2.82x fewer particles for Cytosponge procedures (*p*<0.05). Cytosponge removal is not significant (*p*=0.166) nor is application of throat spray (*p=*0.438) compared to OGD, bearing in mind throat spray is usually not required for Cytosponge. In the droplet size range we find that the application of throat spray produces 9.8x fewer particles (*p*<0.05) for Cytosponge procedures compared with OGD. Cytosponge removal is not significantly different between the two procedure types for droplets (*p=*0.255) nor is coughing/gagging (*p*=0.282).

## Discussion

We find that, over the entire length of the procedure, Cytosponge procedures produce significantly less aerosols (raw data: 2.16x, background subtracted: 4.39x) than OGD procedures, an effect comparable to replacing OGD with trans-nasal procedures (raw data: 2.00x). Our event-based analysis suggests throat spray is a major source of aerosols, similar to OGD, but we observe a reduction in droplet size particles. This may be due to the seated upright position of the patient causing more particles to fall to the floor before reaching the detector. However, throat spray is only used 22% of the time in our observed Cytosponge procedures and only 5-10% of the time for Cytosponge procedures generally, compared to 100% of the time in our observed OGD procedures. Our analysis of Cytosponge procedures with no throat spray does not show a significant reduction in aerosols (2.08x vs OGD), suggesting that throat-spray contributions are largely transient and do not contribute significantly to total particles measured over the entire procedure length.

Coughing/gagging in Cytosponge procedures occurs with similar frequency to OGD but produces significantly less aerosols, which may be due to the different patient position, lack of insufflation, lack of water spraying, and the mouth being mostly closed. Reduction in aerosols, which can stay airborne for hours, reduces infection transmission risk from coughing/gagging. The removal of the Cytosponge is comparable to the extubation of an endoscope in particle quantity and size, likely due to the similar mechanical forces in both cases.

In future studies a larger sample size could be used to increase statistical confidence, particularly against large variations in background particle levels, though our background subtraction method goes some way towards this. Cytosponge procedures should be recorded in a wider range of rooms, to examine the effect of room sizes and ventilation, and administered by numerous different medical staff to examine the effect of procedure technique. A larger sample size would also enable analysis of the impact of variables (age, BMI, smoking etc) on particle production, enabling triage for risk mitigation.

These data suggest that Cytosponge is a lower risk for aerosol generation compared with OGD especially when throat spray is not required. In light of this use of throat spray prior to the removal of Cytosponge, which takes place over a few seconds, should be discouraged especially during periods of high risk for respiratory virus transmission. Use of office based, non-endoscopic procedures such as Cytosponge have a number of advantages including ease of access and administration, lower costs, high patient acceptability; and we can now add lower risk of aerosol generation.

## Data Availability

Data associated with this publication is available at http://dx.doi.org/10.17639/nott.XXX Code used for data analysis in this publication can be found at https://github.com/gsdgordon/aerosols

## Notes

**CONFLICTS OF INTERESTS** RCF is named on patents related to Cytosponge and assays which have been licensed by the Medical Research Council to Covidien (now Medtronic). RCF is a co-founder and shareholder (<3%) for Cyted Ltd.

### Competing Interest Statement

Rebecca Fitzgerald is named on patents related to Cytosponge and assays which have been licensed by the Medical Research Council to Covidien (now Medtronic). RCF is a co-founder and shareholder (<3%) for Cyted Ltd.

### Funding Statement

The authors thank Norgine Pharmaceuticals for sponsoring the purchase of a particle counter. GSDG acknowledges a UKRI Future Leaders Fellowship (MR/T041951/1).

### Author Declarations

Approval was obtained from Wales Ethics Committee Integrated Research Application System (IRAS) no. 285595 and England Research Ethics Committee IRAS no. 283505, amendment 3.

